# Diversity, Equity, and Inclusion or Bigotry, Inequity, and Exclusion? Applicant Perceptions of Obstetrics and Gynecology Residency Programs

**DOI:** 10.1101/2023.10.13.23297010

**Authors:** Corinne M. Hale, Elise S. Cowley, Shanaya Hebgen, Ryan Spencer, Laura Jacques

## Abstract

**Introduction:** Residency applicants often evidence of Obstetrics and Gynecology (ObGyn) residency programs’ commitments to diversity, equity, and inclusion. Online peer-to-peer discussions suggest applicants evaluate program commitment to diversity, equity, and inclusion through formal and informal channels and share their insights and experiences to help each other form opinions about program culture.

**Methods:** The study team examined applicant descriptions of ObGyn residency programs using qualitative data collected from a public resource available on the social media platform, Reddit. Using an inductive approach, the team analyzed anonymous comments in a shared document called “2020-2021 OB GYN Residency Applicant Spreadsheet” housed within Reddit to better understand applicants’ experiences with and perceptions of US ObGyn residency programs. Codes were collectively determined and assigned to 731 unique comments; this paper focuses on the codes directly and indirectly related to diversity, equity, and inclusion as well as racism, discrimination, and bigotry.

**Results:** We found that applicants used visual, verbal, and behavioral cues to analyze ObGyn residency programs. Students primarily used perceptions gained in program interview days, social events, faculty social media profiles, and program websites to decipher programs’ values and internal commitment to pursuing diversity, equity, and inclusion.

**Conclusions:** This study emphasizes the value applicants place on residency programs’ commitments to DEI. Programs that can clearly demonstrate their dedication to DEI principles are more likely to attract a diverse applicant pool that will become the future medical workforce. Programs can provide informative context related to DEI undertakings, reducing the risk that applicants will draw erroneous conclusions from partial information.

## Introduction

In 2020 and 2021, the US experienced significant social and cultural events that brought issues of social justice and racial inequity to the forefront of public consciousness. The unjust killing of George Floyd set in motion a national recognition of racial injustices and social justice movements, including the Black Lives Matter Movement and White Coats for Black Lives.^1,2^ The COVID-19 pandemic further illuminated racial disparities such as the increased risk of SARS-CoV2 infection, hospitalization, and death among Black and Hispanic populations compared to White non-Hispanic.^3^

Public health data shows that Black, Indigenous, and People of Color continue to have disproportionately worse health outcomes than their White counterparts and are more likely to face barriers to receiving quality healthcare.^4,5^ Research also suggests that racially concordant care has potential to improve patient-physician communication and reduce medical costs for ethnic minority patients.^6,7^ Residency programs are training future physicians to enter into the workforce; building a diverse applicant and trainee pool grounded in cultural humility is critical to supporting the increasingly diverse US patient population.

We turned to a publicly accessible forum on Reddit, a popular social media site, to better understand residency applicants’ perceptions of DEI prioritization within residency programs during the 2020-2021 Obstetrics and Gynecology (ObGyn) application cycle. Given the heightened attention to racism in the US in 2020-2021, we anticipated that heightened awareness of DEI-related issues would provide a robust sample of material to analyze.

## Methods

In this study, we examined applicant perceptions of ObGyn residency programs using data collected from a public resource available on the social media platform, Reddit.

Reddit is a social-media platform for text-based information sharing and discussion among anonymous users on a wide variety of topics. With over 52 million daily active users, Reddit is a popular social media platform in the US with average age of users between 22 and 34 years.^8,9^ Users curate their own feeds by selecting “subreddits” to follow or participate in. The site aggregates records of each subreddit and makes them publicly available in Google spreadsheets. Reddit is increasingly used as a qualitative data source among researchers from various disciplines, including the medical sciences, social sciences, and the humanities.^10^ We analyzed comments within one of these shared documents to better understand applicants’ experiences with and perceptions of US ObGyn residency programs We accessed the “2020-21 OB GYN Residency Applicant Spreadsheet” Google spreadsheet posted to the 2020-2021 Residency Interview Spreadsheet megathread on the r/medicalschool subreddit on May 27, 2022. Each spreadsheet houses 12 months of commentary in approximately 10 different tabs of content accessible on the public domain.

We analyzed comments in three of the 10 tabs from the spreadsheet - “Name & Shame,” “Student Reviews,” and “PM_Pearls.” Students use these particular tabs to share program-specific experiences and perceptions and ask questions in a conversational style where qualitative methods can be easily applied. The research team removed all identifying information from posts, including usernames and location, and removed duplicate posts. We imported 731 unique anonymous posts to NVivo 12(QSR International, Burlington, MA), a qualitative analysis software program.

We used thematic analysis to analyze poster comments. We used an inductive approach to generate codes. Three study team members independently reviewed a randomly assigned subset of comments and identified possible codes. Then, as a group the researchers compared independently identified codes and adjusted codes until consensus was reached. The final codebook contained 11 codes and 38 subcodes. Three investigators (CH, ESC, and SH) coded posts within NVivo together to reach consistency in code application, then coded the remaining posts. All five authors independently read the coding reports, met as a group to discuss and refine, and then came to a consensus on themes.

This paper focuses on our “Diversity, Equity, and Inclusion” code and related subcodes. We follow the American Psychological Association framework of diversity, equity, and inclusion as pursuing fair treatment and representation for all people, particularly people who have been historically underrepresented in certain spaces.^11^ We coded any comment related to DEI or lack of DEI within this code: comments on program initiatives on diversity, equity, and inclusion; perceived faculty, resident, and applicant demographic make-up; applicant understanding of program DEI initiatives; and mention of program responses to DEI-related questions. Over three fourths of the 80 comments coded under DEI included follow-up comments, endorsements, or refutations indicating applicants are keenly interested in DEI issues.

IRB approval was waived for this project by the University of Wisconsin-Madison reviewers because the researchers used publicly available data.

## Results

We found that ObGyn residency applicants used visual, verbal, and behavioral cues to assess how and whether programs prioritized diversity, equity, and inclusion at the resident, faculty, and program level. Students primarily used perceptions gained from program interview days, social events, faculty social media profiles, and program websites to decipher programs’ values and internal commitment to pursuing diversity, equity, and inclusion. We found that applicants relied on: 1) Visual representation of diversity, 2) Verbal communication around DEI, and 3) Interviewer actions to better understand DEI prioritization within residency programs. We use exemplar posts to illustrate themes and have preserved the wording, punctuation, grammar, and syntax of the original text.

### Theme 1: Visual Representation of Diversity

ObGyn residency interview days moved to virtual platforms for the 2020-2021 application cycle. According to the commentary we analyzed, applicants found themselves turning to different resources to get a sense of a program’s diversity. Applicants seemed to peruse program websites to view faculty and current resident bios and relied on faculty, current resident, and applicant appearances during interviews, meet and greets, and social events to gauge diversity.

One applicant commented, *“[They] had us attend a six-hour diversity second look*^*1*^ *with ZERO Black or Latinx (from appearance and names) applicants present. Idk what they’re doing but that was my sign*.” This comment may imply a potential connection between how applicants use demographic make-up to discern how residency programs prioritize DEI based on their visual perceptions of who the program invites to interviews.

Other posters stated similar concerns about the same program, “*Was disappointed by lack of diversity in applicant pool during my IV [interview] day*” followed up by responses “*No Black applicants on my IV [interview] day*” and then “*They have ZERO black residents right now*.” Applicants called out racial homogeneity at other residency programs in comments like “*New intern class (of 2025) is completely white*” followed by “*they have very little diversity in the current residents as well*.”

### Theme 2: Verbal Communication around DEI

Reflecting on their interview days, applicants highlighted racist comments from program representatives including program chairs, directors, faculty, and residents. When evaluating verbal communications related to DEI, we identified three subthemes from posters’ comments: 1) Inadequate Responses to Applicant DEI Questions, 2) Bigotry, Racism, and Microaggressions, and 3) DEI Gone Wrong.

#### Inadequate Responses to Applicant DEI Questions

Applicants often listed program responses to DEI-related questions posed by applicants as program-specific “shames.” Here we define “shames” as issues, comments, or activities enacted by programs or program representatives that caused applicants distress or frustration.

Given the sociopolitical context of 2020-2021, applicants may have been more prepared to pose questions related to DEI initiatives within residency programs than in prior years. However, program representative responses to DEI-related questions varied and revealed problematic understandings of what diversity can and should look like. Applicants described many responses to their DEI inquiries as inadequate, and some as sexist, racist, or simply clueless.

One applicant mentioned “*Question on diversity efforts was met with how they look for ‘competent’ applicants*.” Another stated they “*asked about [DEI] efforts and was told there were none within the department*.” And yet another commenter shared “*No DEI efforts…chair said their DEI initiatives are ‘their patients’*.” These reflections may suggest that DEI initiatives are not a priority and program representatives are not prepared to respond.

Other responses reinforced stereotypes rather than undermining them. Take the following comment related to sexuality:

> “*An applicant asked ‘How many LGBTQ+ faculty do you have in the department?’ The resident’s response was something like: ‘I can’t think of any off the top of my head. But our new fellow who’s starting next year looks super gay*.*’ It felt a little tone-deaf*.*”*

Discrimination in medicine can be based on perceived racial or ethnic background, gender, sexuality, class, ability, or other factors. These factors intersect for many applicants. Yet some program comments seemed to erase these intersections, revealing to applicants a misconception about what DEI can mean.

> “*During the social a WOC [woman of color] applicant asked about diversity initiatives and the residents started their answer talking about the male-female breakdown of their program*.” Followed by a response “*I was in this interview and almost died when the WOC applicant asked about diversity and they proceeded to brag about having a lot of MEN in the program…big yikes*.”

#### Bigotry, Racism, and Microaggressions

The Reddit commentary suggests residency applicants use verbal cues to evaluate perceived prejudices of program representatives that show up in both subtle and blatant ways. According to our analyses, prejudices, racism, and discrimination seem to manifest in conversations during the informal aspects of residency interview days and social events.

In one instance, a commenter recalled “*Faculty member made a super concerning racist remark regarding current residents*.” Additional examples pinpoint the fact that applicants are paying close attention to people in power such as program directors, associate directors, and program chairs.

Commenters provided a number of cases where program leadership used inappropriate and harmful language concerning migrants: “*Program chair repeatedly referred to undocumented as ‘illegal immigrants*” and “*Chair used the words ‘undocumented aliens*.”

While we do not know the context that prompted these comments and others like it, applicants appeared to be paying attention to what program representatives say and how they say it. We found this is to be the case not only for faculty, but current residents as well. “*In pre-interview event, resident made several borderline derogatory remarks about the patient population*,” one applicant recollected.

In a separate example, an applicant shared “*A resident stated ‘I don’t live in [City] proper, because you know, it’s hood,’ which was just absolutely appalling and so blatantly racist especially given the patient population that this hospital serves*.” Followed by another comment about the same program “*Resident also called [City] the ‘hood’ and stated not many residents live there because ‘even though it’s convenient to live in [City], it’s more convenient to be alive during residency*.’ *And lastly, during faculty panel interview, faculty member who knew my family is from Africa asked if I was Rastafarian (no) and then followed up with a quiz history question about my family’s home country. Very off-putting*.”

This commentary reflects microaggressions presented by both faculty and current residents. Our analysis indicates that these microaggressions, or usage of derogatory language or actions directed toward marginalized groups of people, were often pointed at and impacted applicants themselves or the patient population the program serves.^12^ Applicants of color seemed to be subjected to inappropriate and illegal questioning from program representatives on the basis of their perceived “difference.”

> *“[I] kept getting pressured to explain why I wouldn’t return to ‘my home country’ that I haven’t lived in for over a decade, and when my first language is English, for training or after residency; ‘I just don’t know how you’re going to get through residency when your family lives in X City’ [statement from the program director] (a city I’ve never been to and have zero connections to, but with a high population of people with my ethnicity). Of course, [I] got the ‘Your English is so good’ thrown in by half my interviewers. Just a ton of microaggressions I have neither the energy or interest in repeating. I thought they were nice and a great group when I attended their virtual meet and greets, but none of that was apparent one-on-one and gave the impression that the vibe only includes you if you’re the right color/ethnicity. Not saying this happens to everybody, but this was my experience*.”

If applicants were to take the experiences shared in the posts as truth, it would appear that international students were at a particular disadvantage and subject to potential discrimination. The following thread elucidates this possibility.

> “*Diversity person stated in URM meet and greet that they ‘try to interview 30% URM however they are ‘grateful’ to be a top program and not have to interview IMGS [International Medical Graduates]*.”
>
> Then, “*It’s so sad that they think this is a good effort for inclusion*.”

Followed by “*wait, they said they were GRATEFUL not to have to interview IMGs [International Medical Graduates]?!?! I’m sorry, but I am an MD student who has worked with IMG [International Medical Graduate] residents and they are some of the most qualified, brilliant, hardworking, incredible physicians I know. How dare anyone act like interviewing international graduates is a burden. They bring so much to a training program and this point of view is so disappointing*.”

In addition to racist and xenophobic encounters, we also reviewed comments related to instances of ableism where program representatives were discriminating against people with disabilities and students who requested testing accommodations. There were not many comments related to disability, but we acknowledge the importance of the topic for furthering DEI conversations.

#### DEI Gone Wrong

Prejudices can also be veiled under the guise of building equity. Even in efforts to promote and support diversity, equity, and inclusion, programs may still fall short according to applicant interpretations. The following example suggest that applicants may hold their own ideas and presumptions about how program DEI efforts and initiatives should and should not be communicated.

> “*There is this fellow that is on the 2021-2022 admissions committee. She straight up said she is looking for brown applicants that have an extreme leftist mentality and are able to openly speak about controversial topics in medicine during the interview. She said if you don’t have the same mindset as “us” (aka herself) then I am not interested in having you attend my program. Was mentioned she is not looking for southern applicants as they are ‘too proper*.*’ Also asked me to describe racism to her during the interview. I would rather not attend a program with such a malignant fellow representing the school*.*”*

In a possible attempt to achieve equality, an associate program director asked an applicant of color what they “*do to be anti-racist*” in an interview. The applicant posted “*I’m Black so that was super off putting and it was a very poor question to ask a Black woman*.”

Self-ascribed underrepresented applicants proffered experiences where interviewers were seemingly trying to offer encouragement, but their sentiments belied potential notions of diversity quotas over applicant competency.

> “*I was asked what other programs I am applying to. I was also told, and this was super cringe, that if there are two applicants and I was a worse candidate (he did hand gestures) I would get picked as an URM just because of that which was weird. But also, he said there is a grant for programs who take URM residents so it would be a benefit to them. For a program that is super diverse that did not feel good*.”
>
> And “*I was told the same thing and agree, it was not a nice thing to hear*.*”*

### Theme 3: Actions Speak Louder than Words

When programs are not proactively addressing diversity, pursuing equitable practices or communicating on the subject, applicants are forced to fill in the blanks by monitoring the actions of program representatives. We reference actions to demarcate the unspoken behaviors, or signals, applicants observe about programs and program representatives.

Applicants referenced behavioral observations as cues to discern program commitment to DEI and overall culture. These observations extend to social media profiles. One applicant shared “*The chair of the department posts insane MAGA (make American great again) stuff on his [Facebook] (one comment being about shooting liberals) and talks about how abortion is immoral*.” Given the public nature of social media outlets, personal content shared is easily accessible. Comments like this example may flag applicant concerns and warrant program attention, especially if program leaders’ philosophies do not align with program or institutional values.

Other instances were indicative of problematic internal affairs that suggested privilege and power work to protect people in leadership positions. One poster referenced that an associate dean of graduate medical education had been sued for discrimination at least three times, but has maintained their academic position. Another applicant referenced a pending 2021 lawsuit filed against a dean regarding “discrimination and racism.” Applicants seem to lean on these types of situations as barometers for program culture, including DEI commitment, and to build awareness among peers.

Even when DEI efforts are being made, applicants are also concerned with who takes the lead on program initiatives. As one person stated, “*strong momentum for DEI advocacy yet the only faculty members leading the DEI initiatives are junior faculty/recent graduates*.” Without evident senior faculty buy-in and engagement, applicants may infer that DEI initiatives are not a program priority.

## Discussion

The lack of racial and ethnic diversity within the US physician workforce has been a persistent and recognized issue.^13^ Residency selection plays a critical role in determining the diversity and representation within medical specialties, providing pivotal opportunities to address disparities.^14^ Representation varies significantly across medical specialties; Obstetrics and Gynecology residency has a higher proportion of underrepresented in medicine (URM) applicants and matriculants than other medical specialties.^15^ However, despite an Accreditation Council for Graduate Medical Education mandate and discipline specific efforts,^16,17^ the percentage of URM residents has remained stable over time and still falls short of reflecting the ethnic and racial diversity within the US population.^18^ This lack of representation at the residency level influences the future workforce caring for an increasingly diversified US patient population.

It is imperative for programs to understand how they are communicating their commitment to DEI to attract applicants, meet the ACGME requirement, and train future-physicians whose demographics more accurately reflect the US population. One opportunity to garner insights on how programs are perceived includes inviting feedback from applicants themselves. Our research addresses this major gap in the literature by elevating applicant voices. Much of the literature available on DEI efforts within graduate medical education keys in on how program leaders, including chairs and directors, as well as faculty, understand diversity, equity and inclusion measures or needs within their own institutions.^19^ However, these studies lack critical voices: they generally do not include applicant or trainee narratives. We found that applicants discuss racism, bigotry, and discrimination within programs in ways that could inform program improvement efforts. Applicants are paying close attention to sources including program websites, faculty members’ personal social media pages, and interview spaces to draw conclusions about program prioritization of DEI. Insights gleaned from ObGyn applicant experiences can help inform future DEI-related practices in graduate medical education.

It behooves ObGyn residency programs to have clear lines of communication on how programs are addressing social issues and questions of diversity, equity, and inclusion. Otherwise, applicants will construct images of programs based on their own observations and perceptions. These perceptions are entangled with who is in power in the program, the demographic make-up of faculty, staff, and trainees, as well as how program representatives present themselves as stewards of the program through their communication and behavior.

The strengths of this study include the use of Reddit as a data source that affords users anonymity, which may limit social desirability bias by creating a safe space for users to openly share experiences without fear of personal consequences. We analyzed a high volume of comments and the richness of the data set allowed us to reach thematic saturation.

Reddit is a popular and commonly used website among people in their twenties and thirties, prime ages for applying for residency. The Reddit spreadsheet forum allowed us to see how users engaged with each other by interacting with posts. We were able to see discussions between peers which included exclamations of solidarity and affirmations as well as disagreements. Comments came directly from applicants and were not guided by researcher agendas or bias, organically reflecting what is important to students. Additionally, applicants nationwide can utilize the Reddit forum to discuss any ObGyn residency program in the country, not just a single institution. The national applicability allowed us to get a good pulse on what applicants think broadly and may deem important when applying for residency. Nevertheless, more research is necessary to further understand the depth and scope of DEI considerations as drivers of choice for ObGyn residency applicant program decision-making.

Our analysis was limited to relying on data saved in the ObGyn spreadsheet eliminating the possibility of follow-up or eliciting further feedback from commenters for clarification or more details. We relied on anonymous comments voluntarily shared, but more research directly asking medical students and trainees about their experiences and concerns related to programs’ commitment to DEI tenants is necessary. Future research could highlight student and applicant narratives collected through in-depth interviews to better understand how applicants perceive programs and how these perceptions influence their decision-making processes on where to attend residency.

## Conclusions

Applicants to obstetrics and gynecology residency training programs assess those programs’ commitments to diversity, equity, and inclusion on a range of grounds—and discuss their perceptions and conclusions with their peers. We found that applicants are searching for demographic diversity within programs at all levels, equitable selection processes and training practices, and inclusive environments to learn and attend to patients. Residency programs can benefit from understanding how their communication practices and program representatives contribute to applicant perceptions of program advancement of DEI principles.

## Data Availability

All data produced in the present study are available upon reasonable request to the authors.

## Author Notes

This project was supported by funding from the University of Wisconsin-Madison Department of Obstetrics and Gynecology.

Elements of this work were previously presented at the APGO Martin L. Stone, MD, Faculty Development Seminar (FDS) on January 10th, 2023 in Scottsdale, Arizona.

None of the authors have a financial or other conflict of interest.

The authors have no disclaimers.

## Footnotes

1 A “second look” is an opportunity for interested applicants to participate in a virtual follow-up visit with the residency program. Second looks are offered after first interviews are complete but before the Match.

## Notes

### Competing Interest Statement

The authors have declared no competing interest.

### Author Declarations

The Institutional Review Board of the University of Wisconsin-Madison waived ethical approval for this work because the data was publicly available on the social media platform, Reddit.

